# Impact of body mass index on 5-year survival rates in patients undergoing allogeneic hematopoietic stem cell transplantation

**DOI:** 10.1101/2022.02.25.22269995

**Authors:** Takashi Aoyama, Akifumi Notsu, Koki Ichimaru, Masanori Tsuji, Kanako Yoshitsugu, Masafumi Fukaya, Terukazu Enami, Takashi Ikeda

## Abstract

**Background:** The association between cancer survival and body mass index (BMI) has been elucidated. However, the impact of patients’ baseline characteristics on allogeneic hematopoietic stem cell transplantation (allo-HSCT) outcomes remains unclear.

**Objectives:** To examine the baseline clinical factors associated with 5-year survival rates in patients undergoing allo-HSCT.

**Study design:** This was a retrospective exploratory observational study. Patients (n=113, average age: 55 years; 52 women) who underwent allo-HSCT at the Division of Hematology and Stem Cell Transplantation, Shizuoka Cancer Center, between January 2008 and March 2015 were included in this study.

**Results:** The 5-year survival rate (65%) was associated with the baseline geriatric nutritional risk index (GNRI; odds ratio [OR]=1.20, 95% confidence interval [CI]: 1.03–1.36, P <0.01) and the BMI (OR=1.06, 95% CI: 1.02–1.12, P <0.01). The cut-off values for BMI and GNRI were 20.5 kg/m^2^ and 101 points, respectively (area under the curve, 0.65017 and 0.67637, respectively). The 5-year survival rate was poorer for patients with sarcopenia (41.5%) than for those without sarcopenia prior to allo-HSCT (p=0.05).

**Conclusions:** BMI and GNRI values before allo-HCST pre-treatment were independent predictors of the 5-year survival rates. Patients undergoing allo-HSCT may require nutritional interventions during pre-treatment to reduce the risk of sarcopenia, which affects their survival rates.

## Introduction

A cohort study found that standardized body mass index (BMI) is associated with cancer survival [1]. In Japan, the 5-year survival rates (2009–2011) associated with leukemia, multiple myelomas, and malignant lymphomas have been reported to be 44.0%, 42.8%, and 67.5%, respectively [2]. Allogeneic hematopoietic stem cell transplantation (allo-HSCT) is used to achieve remission in cases of hematopoietic tumors. Body weight loss has been associated with allo-HSCT outcomes [3]; however, this association has not been examined in a multivariate study. Furthermore, no previous study has examined the impact of nutritional assessment on allo-HSCT outcomes. At the Shizuoka Cancer Center (SCC) Division of Hematology and Stem Cell Transplantation, patients are provided with a nutritional intervention using a nutritional pathway [4]; however, the relationships between the patients’ clinical characteristics and outcomes have not yet been examined. The aim of this study was to examine the impact of the patients’ baseline clinical characteristics on allo-HSCT outcomes and to examine the role of nutritional interventions in this context.

## Materials and Methods

This retrospective, exploratory, observational study included 120 patients (age: 16–70 years) who underwent allo-HSCT (transplantation day: day 0) and for whom a nutritional pathway intervention was recommended at the SCC Division of Hematology and Stem Cell Transplantation between January 2008 and March 2015. Patients were excluded from this study if data on their clinical characteristics were lacking. Allo-HSCT pre-treatments included myeloablative conditioning (MAC), reduced-intensity conditioning (RIC), and human leukocyte antigen (HLA)-matching. Transplantation types included unrelated donor bone marrow transplantation (UR-BMT), cord blood transplantation (CBT), and allogeneic peripheral blood stem cell transplantation (allo-PBSCT) [5].

Patients were evaluated before undergoing pre-treatment (T1) and 5 years after receiving their transplant (T2). Remission status at T1 was not evaluated for patients with myelodysplastic syndromes [5] for which the remission status could not be determined. Data were collected on the following variables: 5-year survival rates (using day 0 as the starting point); pre-treatment type (MAC, RIC, and HLA-matching) and transplantation type (UR-BMT, CBT, and allo-PBSCT) and associated survival outcomes; baseline age; BMI; skeletal muscle index (SMI, assessed with a bioelectrical impedance analyzer [BIA: In Body S20®]) [6,7]; skeletal muscle mass (SMM) [8]; fat mass (FM); phase angle [9]; geriatric nutritional risk index (GNRI) [10] values (which were compared between the treatment and transplantation types); hematopoietic cell transplantation-specific comorbidity index (HCT-CI) [11]; and incidence of graft-versus-host disease (GVHD; grade ≥1). The 5-year survival rates were the outcomes of interest. The cut-off values of BMI associated with the outcomes were estimated. The BMI was divided into the following 3 categories: underweight (<18.5 kg/m^2^), normal weight (18.5–24.9 kg/m^2^), and overweight (≥25.0 kg/m^2^) [12]; the 5-year survival rates were compared among these categories. SMI was used to diagnose sarcopenia at T1 (women: <5.7 kg/m^2^, men: <7.0 kg/m^2^) [13]; the survival rates were compared between patients with and without sarcopenia. The association between GNRI was calculated as follows: (14.89 × serum albumin level) + (41.7 × [body weight/ideal body weight]). The association between GNRI at T1 and survival was evaluated; the cut-off value associated with survival outcomes was estimated. Survival rates were compared between groups defined on the basis of a cut-off value of <99 points at T1.

Measurements with the InBody S20® device were taken 2 h after breakfast (from 10:00 a.m. to 12:00 noon). We set the reference extracellular fluid-to-total body fluid ratio as 0.35 and the extracellular water-to-total body water ratio as 0.40. We defined the upper limits of the extracellular fluid-to-total body fluid ratio and the extracellular water-to-body water ratio (indicating mild edema) as 0.35–0.38 and 0.40–0.43, respectively. If edema was noted, the measurements were obtained again to control for the impact of edema. All variables were measured using a high-precision body composition analyzer (InBody S20®); the value frequencies were calculated [14]. Data on the HCT-CI, GVHD, and HLA matching were examined using the Transplant Registry Unified Management Program [15].

### Statistical analyses

Normality of distribution was verified using the Shapiro–Wilk test, and all variables were expressed as median values (minimum–maximum). Univariate logistic regression analysis was used to assess the relationship between the clinical characteristics at T1 and the 5-year survival and GVHD rates. The associations of age with SMI and BMI were assessed using the Pearson’s product moment correlation. The associations of BMI with pre-treatment methods and transplantation types were assessed with an analysis of variance (ANOVA). Cut-off values for the associated factors were calculated using the receiver operating characteristic curve. Differences in the pre-treatment method, HLA, and transplantation types between surviving and non-surviving patients were examined using the chi-square test and the Steel–Dwass test. A survival curve was created using the Kaplan–Meier method, and associations between the survival rate, BMI (3 categories), SMI (sarcopenia), and GNRI were assessed using the log-rank test. All statistical analyses were performed using JMP (version 12.0®; SAS Institute, USA) and R (version 3.6.3; R Core Team [2020], www.r-project.org). Two-sided p-values of <0.05 were considered statistically significant.

### Ethical considerations

This clinical study was performed in accordance with the declaration of Helsinki and approved by the Shizuoka Cancer Center institutional review board (SCC IRB; approval number: J2021-49). Verbal consent was obtained from the patients in accordance with the IRB’s recommendations.

## Results

A total of 113 patients were included in the analysis. Seven patients were excluded due to missing baseline data. The performance status (PS) at T1 was 0 in all cases. The patients’ median age was 55 years (range, 17–70 years; p<0.01 by the Shapiro–Wilk test); the mean ages for women and men were 53 years (17–69 years) and 57 years (17–70 years), respectively (p<0.01). The patients’ characteristics are presented in Table 1.

**Table 1.**
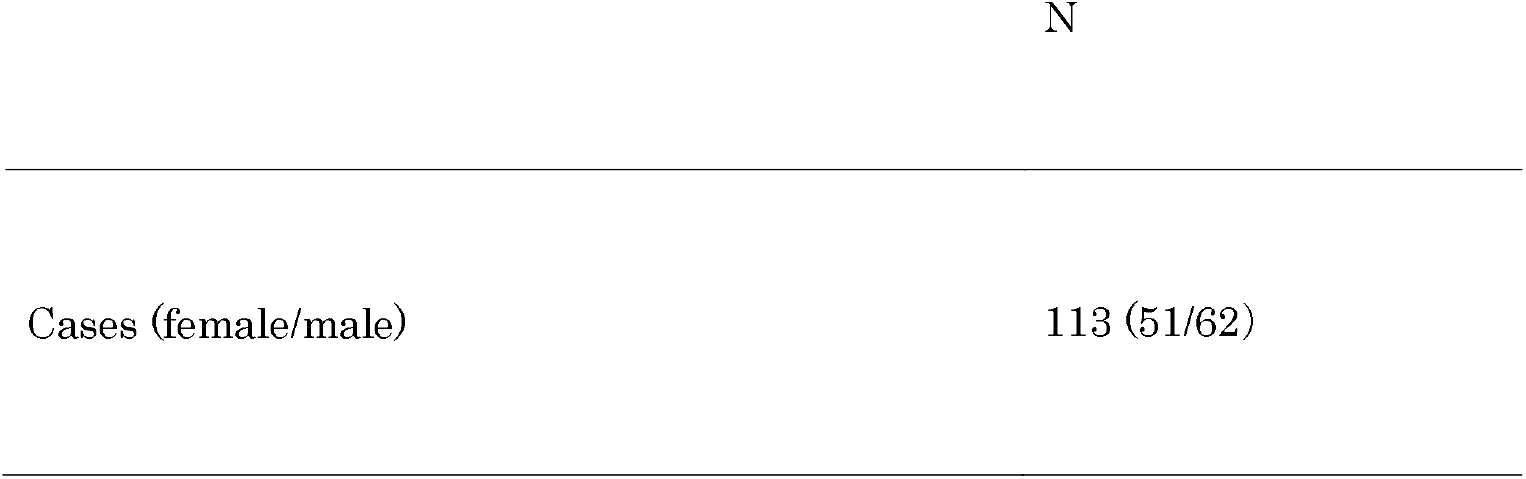
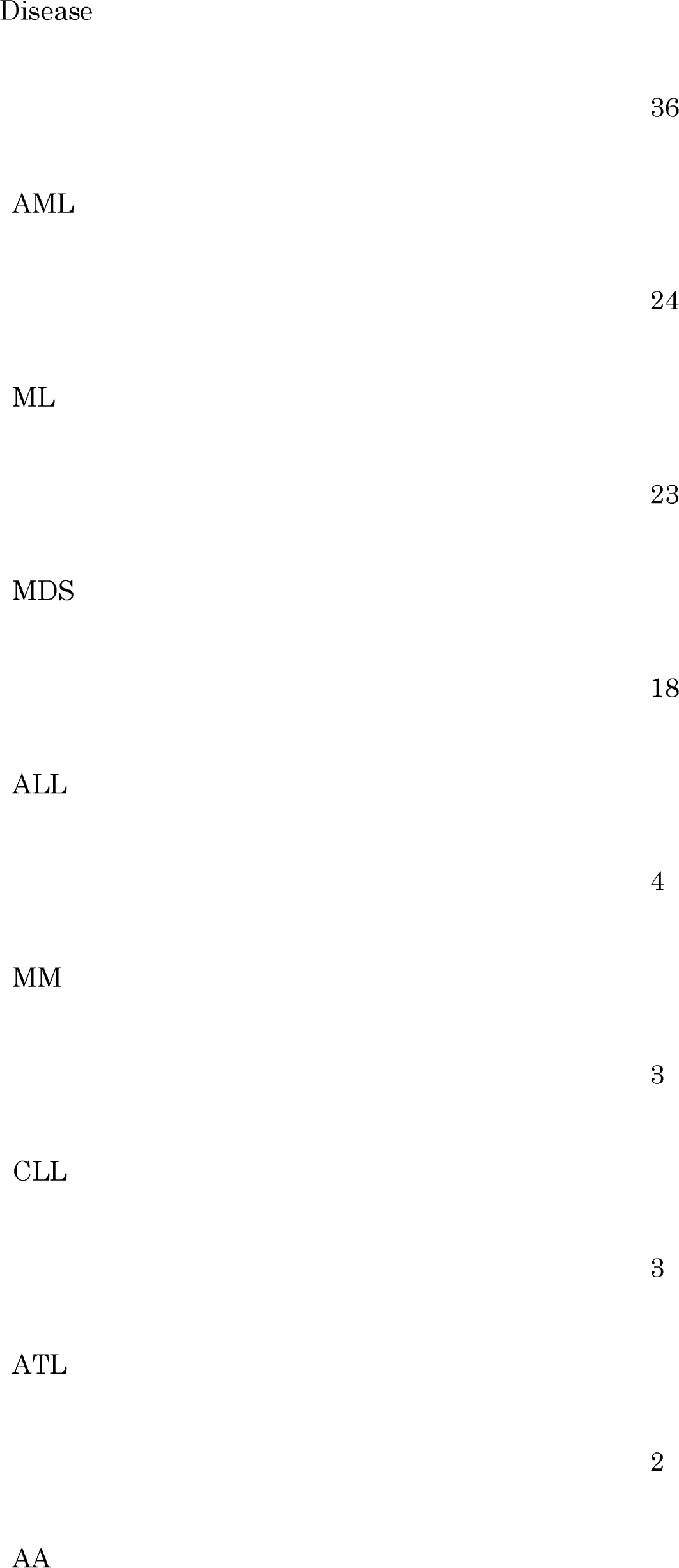
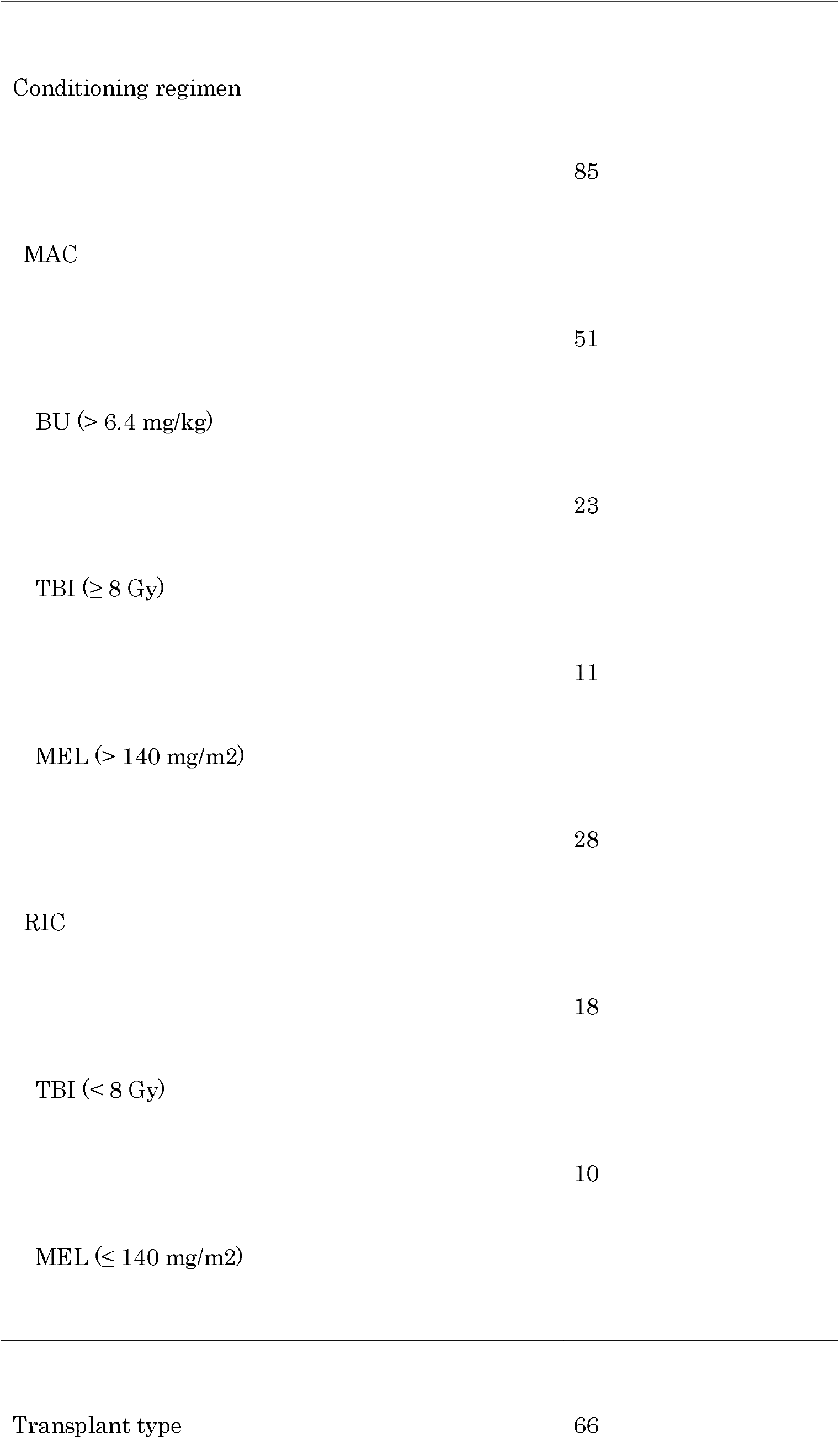
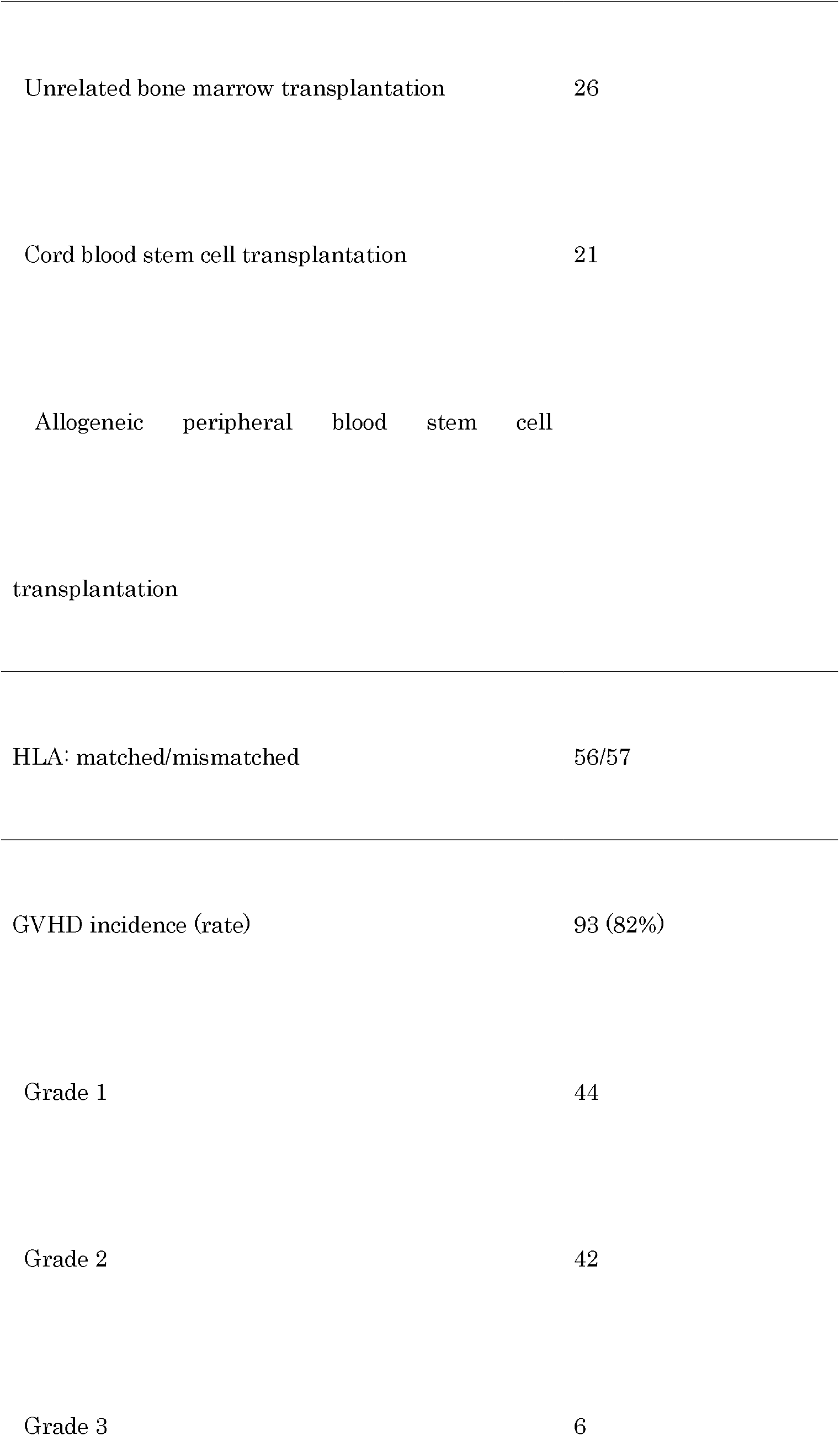
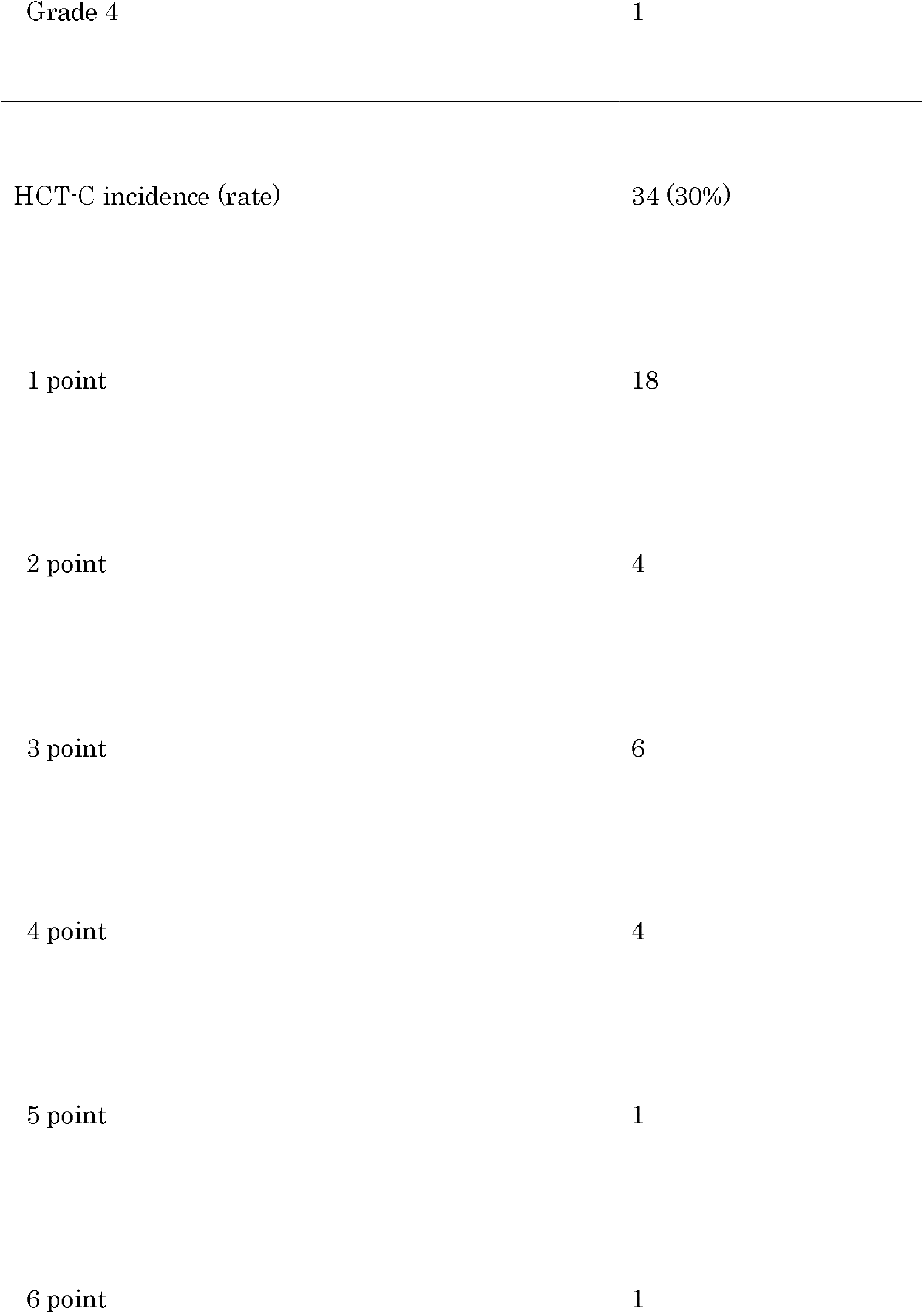
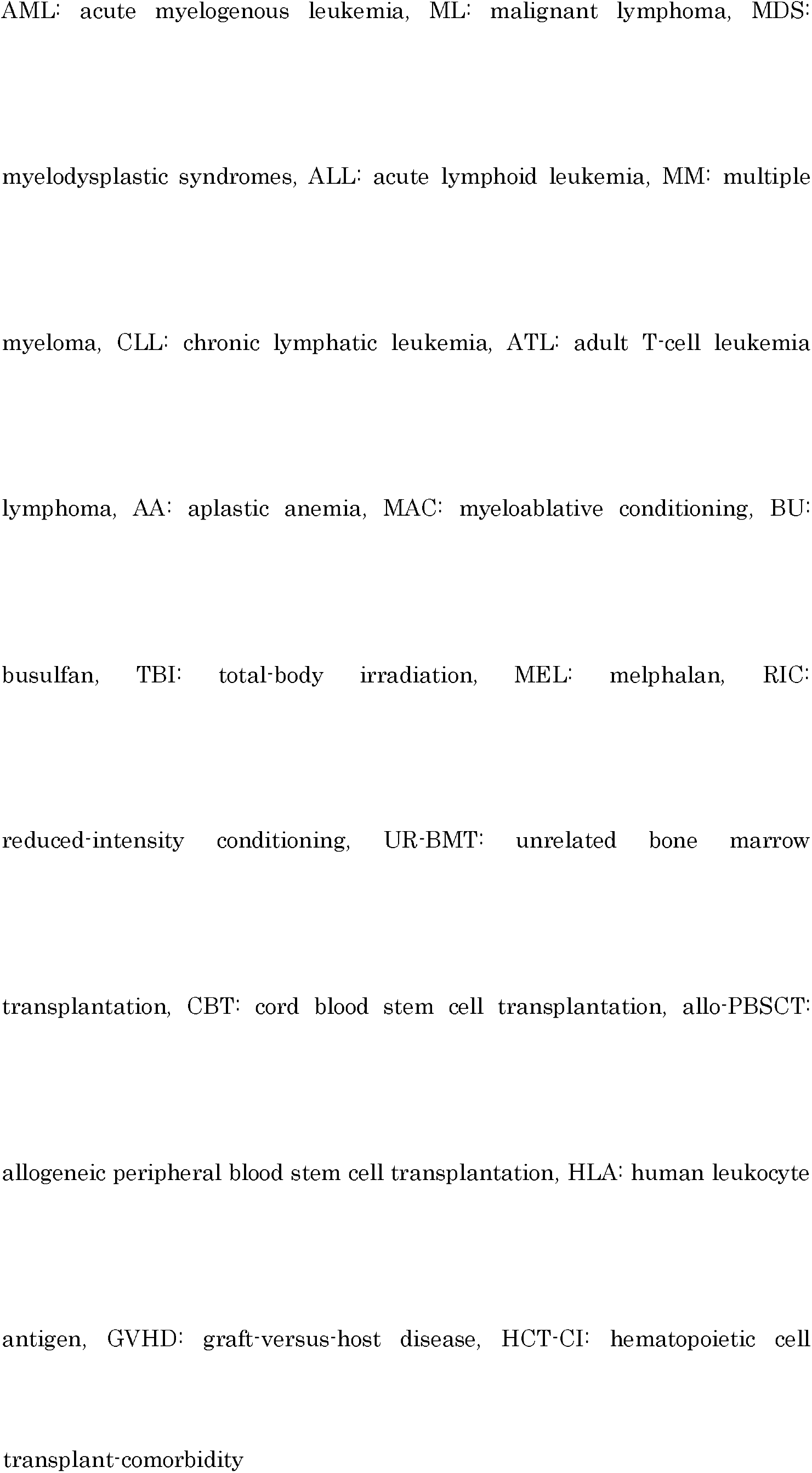
Patients’ background characteristics

The overall 5-year mortality rate was 35% (n=40, including 17 women and 23 men). The non-surviving group included 16 patients with acute myelogenous leukemia (AML; 8 women, 8 men), 7 patients with malignant lymphoma (ML; 3 women, 4 men), 7 patients with myelodysplastic syndromes (MDS; 2 women, 5 men), 6 patients with acute lymphoid leukemia (ALL; 3 women, 3 men), 4 patients with multiple myeloma (MM; 1 woman, 3 men), and 1 patient with chronic myelogenous leukemia (CML; 1 man). The mean age of the non-surviving group was 59 years (range, 18–70 years). At T1, the mean ages of women and men were 54 years (30–64 years) and 61 years (18–70 years), respectively. Furthermore, the overall survival period was 322 days (range, 46–1,756 days; women: 439 days [102–1,496 days], men: 273 days [46–1,756 days]). The overall 5-year survival rate was 65% (73/113; 34/51 women, 39/62 men); these included 20 (9 women, 11 men), 16 (7 women, 9 men), 14 (8 women, 6 men), 15 (6 women, 9 men), 1 (0 women, 1 man), 2 (1 woman, 1 man), 3 (1 woman, 2 men), and 2 (0 women, 2 men) patients diagnosed with AML, ML, MDS, ALL, MM, CML, adult T-cell leukemia (ATL), and aplastic anemia (AA), respectively. A total of 54 and 19 surviving patients underwent MAC and RIC, respectively, at a rate comparable to that observed among non-surviving cases (31 and 9, respectively; p=0.68). Surviving and non-surviving groups had comparable rates of complete and partial HLA matches (37 and 19 in the surviving and 37 and 20 in the non-surviving groups, respectively; p=0.90). Overall, 88, 41, and 24 patients received UR-BMT, CBT, and allo-PBSCT respectively; there were significant differences between the surviving group and the non-surviving group (UR-BMT: 66 vs. 22: p=0.23, CBT: 26 vs. 15: p<.0.01; allo-PBSCT: 21 vs. 3: p<0.05).

There was no association between age and BMI (r=0.04, p=0.69) or between age and SMI (r=0.03, p=0.76) at T1. At T1, the BMI was 21.1 kg/m^2^ in MAC cases (14.1–31.0 kg/m^2^; p=0.55 by the Shapiro–Wilk test) and 21.9 kg/m^2^ in 28 RIC cases (17.8–29.5 kg/m^2^, p=0.07), without any significant between-group differences (ANOVA: p=0.06). Furthermore, at T1, the BMIs were 21.5 kg/m^2^ (14.1–29.3, p=0.99), 22.1 kg/m^2^ (15.0–31.0, p=0.58), and 20.6 kg/m^2^ (16.5–27.5, p=0.91) among patients who received UR-BMT (n=66), CBT (n=26), and allo-PBSCT (n=21), respectively (p=0.18).

Clinical characteristics associated with the onset of GVHD (grade ≥ 1) are presented in Table 2. The BMI, GNRI, SMI (female), SMM (female), and FM (total only) were associated with the 5-year survival rates (Fig. 1).

**Table 2.**
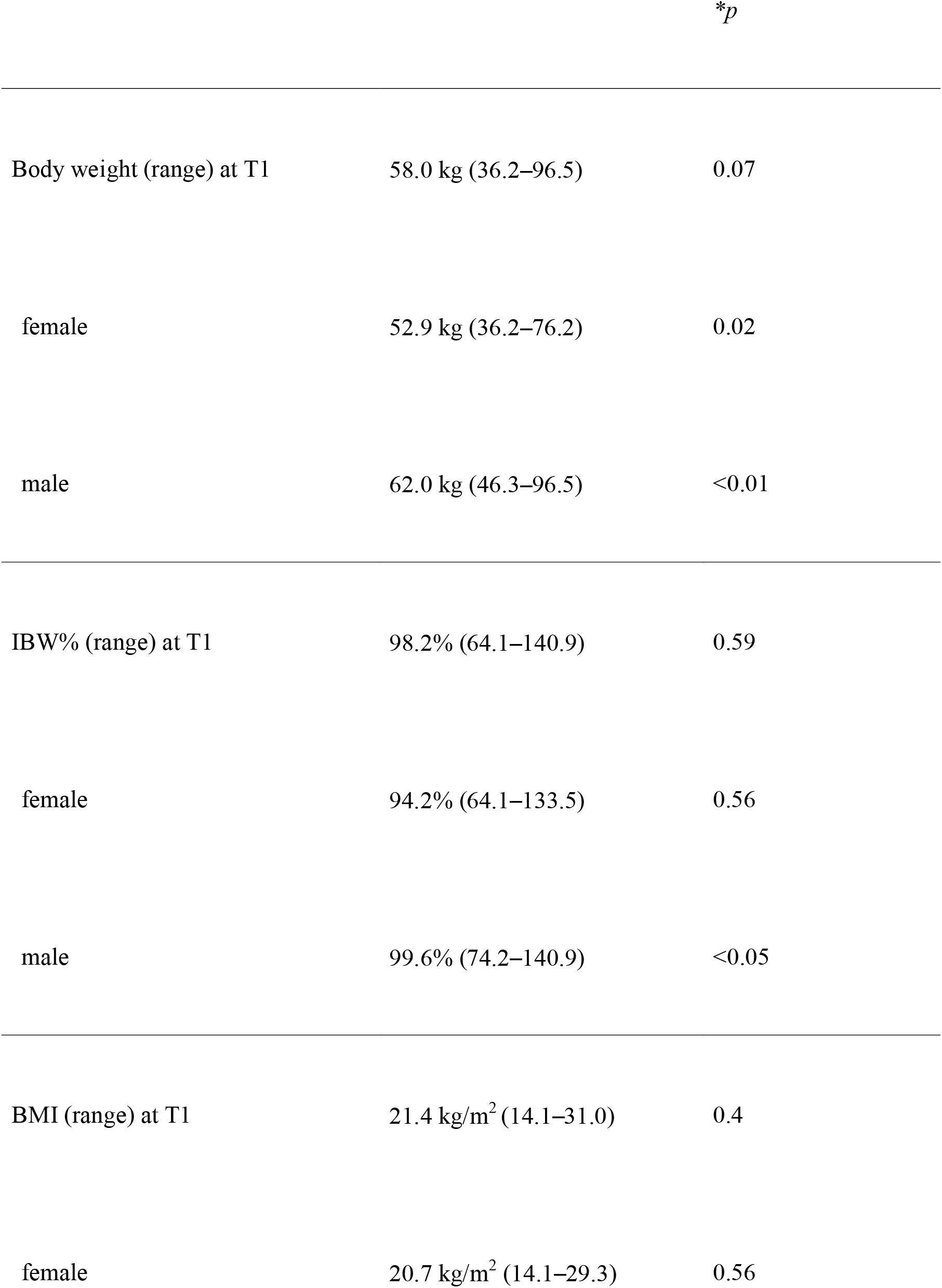
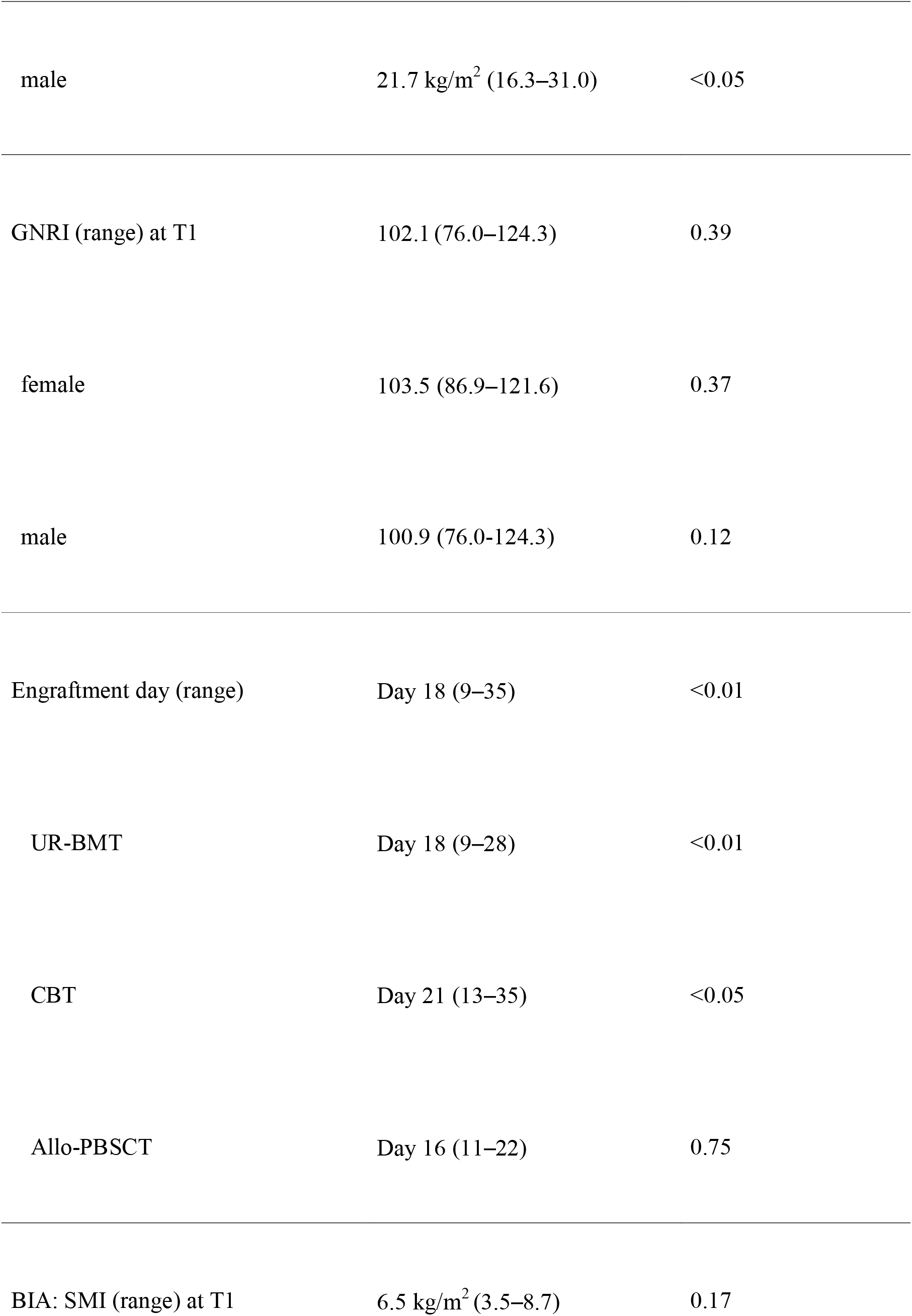
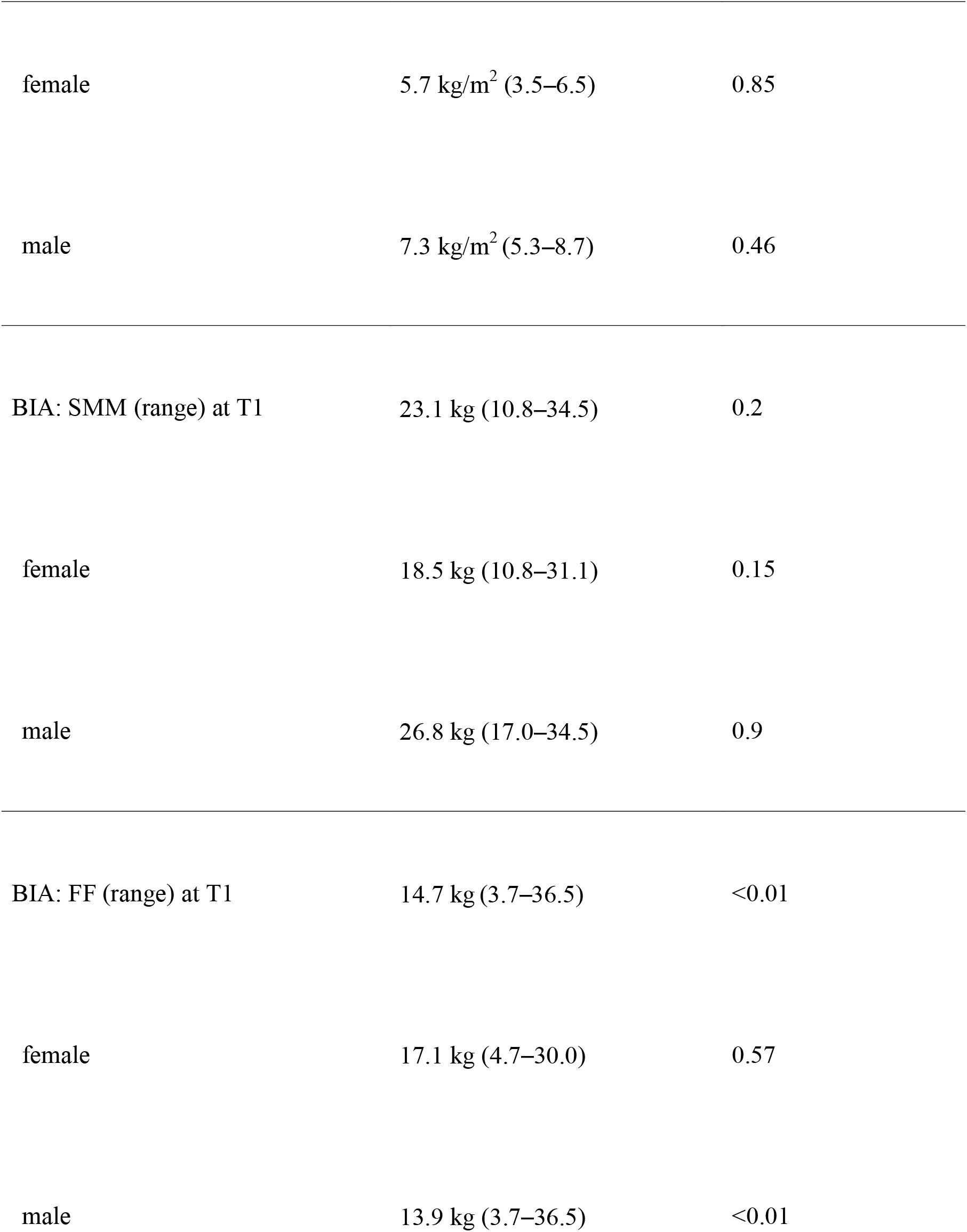
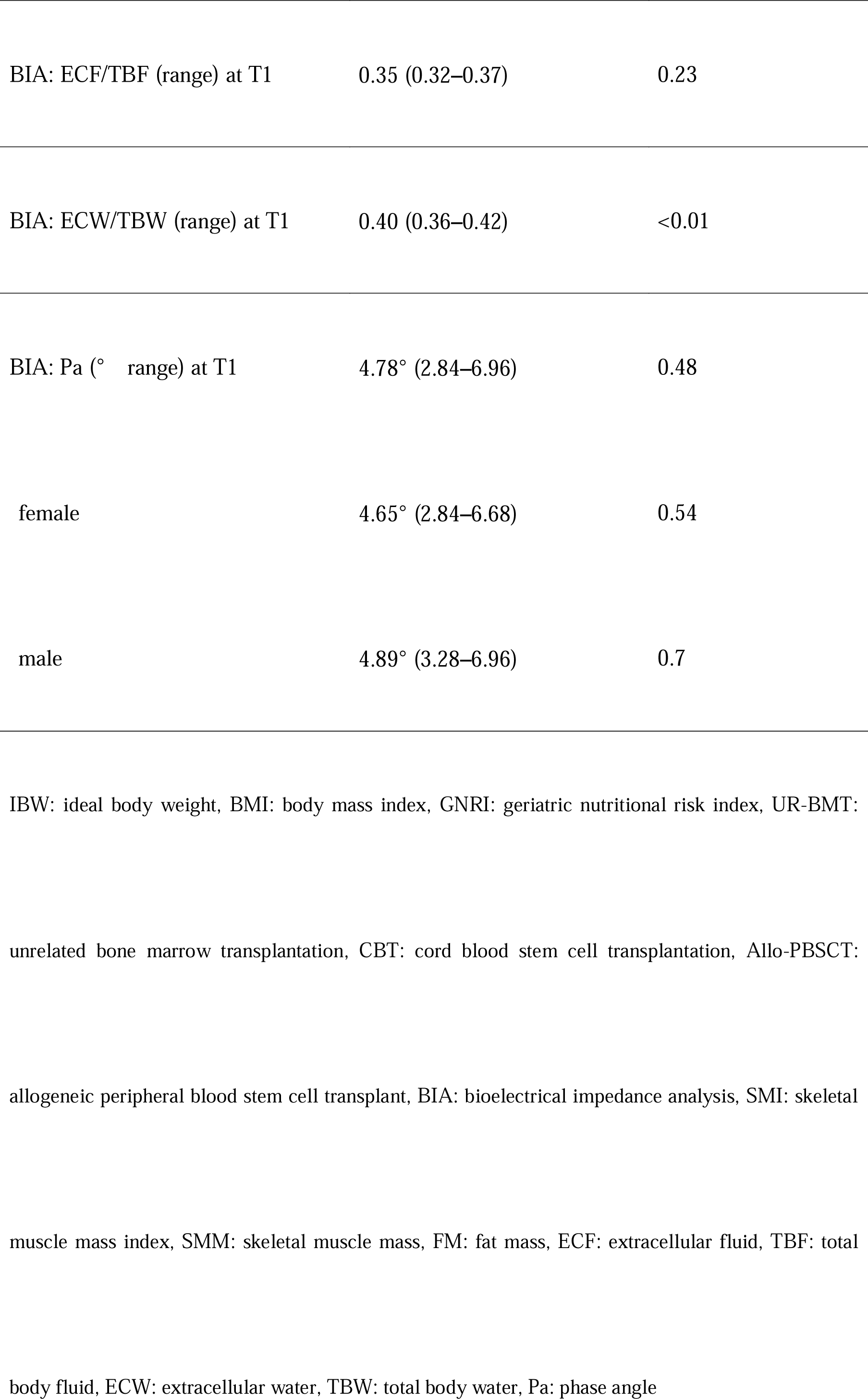
Clinical indicators assessed during the nutritional evaluation

|  |  |  |
| --- | --- | --- |
| Body weight (range) at T1 | 58.0 kg (36.2–96.5) | 0.07 |
| female | 52.9 kg (36.2–76.2) | 0.02 |
| male | 62.0 kg (46.3–96.5) | <0.01 |
| IBW% (range) at T1 | 98.2% (64.1–140.9) | 0.59 |
| female | 94.2% (64.1–133.5) | 0.56 |
| male | 99.6% (74.2–140.9) | <0.05 |
| BMI (range) at T1 | 21.4 kg/m<sup>2</sup> (14.1–31.0) | 0.4 |
| female | 20.7 kg/m<sup>2</sup> (14.1–29.3) | 0.56 |
| male | 21.7 kg/m<sup>2</sup> (16.3–31.0) | <0.05 |
| GNRI (range) at T1 | 102.1 (76.0–124.3) | 0.39 |
| female | 103.5 (86.9–121.6) | 0.37 |
| male | 100.9 (76.0–124.3) | 0.12 |
| Engraftment day (range) | Day 18 (9–35) | <0.01 |
| UR-BMT | Day 18 (9–28) | <0.01 |
| CBT | Day 21 (13–35) | <0.05 |
| Allo-PBSCT | Day 16 (11–22) | 0.75 |
| BIA: SMI (range) at T1 | 6.5 kg/m<sup>2</sup> (3.5–8.7) | 0.17 |
| female | 5.7 kg/m<sup>2</sup> (3.5–6.5) | 0.85 |
| male | 7.3 kg/m<sup>2</sup> (5.3–8.7) | 0.46 |
| BIA: SMM (range) at T1 | 23.1 kg (10.8–34.5) | 0.2 |
| female | 18.5 kg (10.8–31.1) | 0.15 |
| male | 26.8 kg (17.0–34.5) | 0.9 |
| BIA: FF (range) at T1 | 14.7 kg (3.7–36.5) | <0.01 |
| female | 17.1 kg (4.7–30.0) | 0.57 |
| male | 13.9 kg (3.7–36.5) | <0.01 |
| BIA: ECF/TBF (range) at T1 | 0.35 (0.32–0.37) | 0.23 |
| BIA: ECW/TBW (range) at T1 | 0.40 (0.36–0.42) | <0.01 |
| BIA: Pa (° range) at T1 | 4.78° (2.84–6.96) | 0.48 |
| female | 4.65° (2.84–6.68) | 0.54 |
| male | 4.89° (3.28–6.96) | 0.7 |

IBW: ideal body weight, BMI: body mass index, GNRI: geriatric nutritional risk index, UR-BMT:
unrelated bone marrow transplantation, CBT: cord blood stem cell transplantation, Allo-PBSCT:
allogeneic peripheral blood stem cell transplant, BIA: bioelectrical impedance analysis, SMI: skeletal
muscle mass index, SMM: skeletal muscle mass, FM: fat mass, ECF: extracellular fluid, TBF: total
body fluid, ECW: extracellular water, TBW: total body water, Pa: phase angle

**Figure 1.** Odds ratios and confidence intervals associated with baseline clinical characteristics in surviving and non-surviving patients

The BMI cut-off value associated with the 5-year survival rates was 20.5 kg/m^2^ (Fig. 2).

**Figure 2.** Receiver operating characteristics curve for survival, depending on the baseline body mass index

Furthermore, 14, 80, and 19 patients were underweight (<18.5 kg/m^2^), of normal weight (18.5–24.9 kg/m^2^), and overweight (≥25.0 kg/m^2^), respectively. Significant differences were observed in the 5-year survival rates between the underweight, normal weight, and overweight groups (Fig. 3–5). The T1 SMI was indicative of sarcopenia in 47 out of 113 (41.5%) patients. There were differences in the survival rates between patients with and without sarcopenia (Fig. 6).

**Figure 3.** Five-year survival rates by the body mass index category (underweight, normal weight, and overweight)

**Figure 4.** Five-year survival rates by the body mass index category (overweight and underweight)

**Figure 5.** Five-year survival rates by the body mass index category (normal weight and underweight)

**Figure 6.** Five-year survival rate by the sarcopenia status (skeletal muscle index, extremity skeletal muscle mass/tool^2^: women <5.7 kg/m^2^, men <7.0 kg/m^2^)

The GNRI cut-off value associated with the 5-year survival rates was 101 (Fig. 7). The T1 GNRI indicated nutritional risk in 39 out of 113 (34.5%) patients. There was a significant difference in the 5-year survival rates between patients with and without an increased nutritional risk (Fig. 8).

**Figure 7.** Receiver operating characteristic curve for geriatric nutritional risk index values at the baseline and after a 5-year survival

**Figure 8.** Five-year survival rates, stratified by the geriatric nutritional risk index categories (low nutrition: <98)

## Discussion

This study examined the association between the baseline clinical characteristics and 5-year survival rates in patients undergoing allo-HSCT. The overall 5-year survival rate was 65% (mean survival period: 322 days [46–1,756 days]). The survival rates in patients with AML, ALL, CML, and ATL were 36%, 18%, 3% and 3%, respectively, and in those with MM, ML, MDS, and AA were 25%, 67%, 61%, and 100%, respectively. The overall cancer-related and ML-related, leukemia-related, and MM-related survival rates were similar to, higher than, and lower than the corresponding previously reported rates in Japan [2]. Pre-treatment types and HLA matching did not affect the survival rates. In addition, the survival rates were the highest in patients undergoing UR-BMT, followed by in those undergoing allo-PBSCT and CBT. This finding is consistent with that of a previous study [16] and suggests that declining birth rates in Japan may affect donor matching for SCC [17]. In the present study, BMI affected the survival rates at a cut-off value of 20.5 kg/m^2^, which may be a useful reference for nutritional interventions during remission induction therapy or consolidation therapy before allo-HSCT. A significant difference in the 5-year survival rates was observed between patients with low BMIs and patients with normal and high BMIs at baseline; no other associations were observed. These results are consistent with those of a meta-analysis of studies on the impact of BMI on allo-HSCT outcomes [18].

In the present study, GNRI was associated with survival outcomes at a cut-off value of 101, although a previously reported cut-off value (>99) also affected 5-year survival rates. GNRI is an indicator of long-term nutritional status, which may account for the observed association; however, this indicator does not account for differential blood counts, such as lymphocyte levels, which are often affected by hematopoietic tumors. Nevertheless, this finding is consistent with that of a previous study on the impact of BMI on survival. SMI and SMM affected survival rates in women. FM was associated with the overall survival rates; however, no sex-specific associations were observed. Although muscle and FM values were associated with survival in the present study, the mechanisms underlying these associations remain unclear; in fact, the role of the relative metabolic rate in health outcomes remains unclear. Meanwhile, HCT-CI values and GVHD rates were not associated with the survival rates in the present study, suggesting that other pre-treatment and transplant factors may be relevant. Although the T1 PS was 0 in all cases, sarcopenia was confirmed in 41.5% of the patients at T1. To increase the likelihood of a successful allo-HSCT, remission induction therapy and consolidation therapy are used as pre-treatments to reduce the tumor size [5]. In this context, the present results may reflect the limited activities of daily living that patients undergoing pre-treatment may engage in, which may increase the risk of sarcopenia; this in turn warrants a nutritional intervention. In the present study, the baseline BMI and GNRI (both of which depend on body weight) affected survival outcomes of patients undergoing allo-HSCT. The impact of sarcopenia on transplantation outcomes suggests a need for nutritional assessments and interventions before allo-HSCT. This study has some limitations, including the lack of stratification by diagnosis and treatment type and a lack of baseline BIA data (which may have biased the present findings).

In conclusion, the baseline BMI and GNRI values (both of which are associated with body weight), affected the survival outcomes after allo-HSCT, independently of other clinical characteristics. The presence of sarcopenia in patients referred for allo-HSCT suggests a need for nutritional intervention as a part of pre-treatment protocols.

## Data Availability

https://figshare.com/account/home

https://figshare.com/account/home

## Acknowledgements

We would like to thank Editage (www.editage.com) for English language editing. The author would like to thank all dieticians registered at the Shizuoka Cancer Center, Japan, for their assistance with data collection.

## Notes

### Competing Interest Statement

The authors have declared no competing interest.

### Author Declarations

Ethics committee/IRB of Shizuoka Cancer Center gave ethical approval for this work.

